# Predicting Patient Weight from Intracardiac Electrograms: A Study in Electrophysiological Signal Analysis

**DOI:** 10.1101/2024.02.29.24303483

**Authors:** Celal Alagöz

## Abstract

The analysis of electrophysiological signals from the human body has become increasingly crucial, especially given the widespread adoption of wearable technologies and the growing trend of remote and online monitoring. In situations where demographic patient data is unavailable, the evaluation of such information from electrophysiological signals becomes imperative for making well-informed diagnostic and therapeutic decisions, particularly in ambulatory and urgent cases. This study underscores the significance of this necessity by utilizing intracardiac electrograms to predict patient weight.

Intracardiac electrograms were recorded from 44 patients (14 female, with an average age of 59.2±11.5 years) using a 64-pole basket catheter over a duration of 60 seconds. A dataset comprising 2,816 unipolar electrogram signal segments, each lasting 4 seconds, was utilized. Weight, considered as a continuous variable, underwent discretization into k bins with uniformly distributed widths, where various values of k were experimented with. As the value of k increases, class imbalance also increases.

The state-of-the-art time series classification algorithm, Minirocket, was employed alongside the popular machine learning algorithm eXtreme Gradient Boosting (XGBoost). Minirocket consistently demonstrates superior performance compared to XGBoost across all class number scenarios and across all evaluation metrics, such as accuracy, F1 score, and Area Under the Curve (AUC) values, achieving scores of approximately 0.96. Conversely, XGBoost shows signs of overfitting, particularly noticeable in scenarios with higher class imbalance. Tuning probability thresholds for classes could potentially mitigate this issue. Additionally, XGBoost’s performance improves with reduced bin numbers, emphasizing the importance of balanced classes. This study provides novel insights into the predictive capabilities of these algorithms and their implications for personalized medicine and remote health monitoring.

## INTRODUCTION

As technology continues to advance within the healthcare sector, there arises an increasing demand for novel methods to monitor and evaluate patients’ health conditions. In the realm of continuous cardiovascular assessment, wearable and implanted devices, alongside ambulatory monitoring technologies, have become indispensable components (Hong et al., 2019; Sana et al., 2020; Bayoumy et al., 2021). Within this context, the analysis of electrophysiological signals emerges as a valuable tool for assessing patients’ health status. Specifically, the utilization of electrophysiological data such as intracardiac electrograms (EGMs) holds significant potential for evaluating patients’ cardiovascular health and even predicting their demographic information.

Traditionally, patient history is evaluated during the diagnosis phase, aiding in formulating the diagnosis, followed by therapeutic intervention. However, as data collection techniques transition towards remote methods such as Holter monitoring (Zimetbaum and Goldman, 2010), and wearable device usage increases, interpreting signal streams from patients becomes increasingly crucial. Auxiliary information concerning patient demographics significantly influences the development of diagnostic and therapeutic solutions. Yet, in scenarios where patient-specific auxiliary information is unavailable, its assessment becomes imperative.

Patient demographic data encompasses all non-clinical data about a patient, including age, weight, sex, and various other peripheral information. In the context of cardiovascular medicine, correlations between physiological attributes such as age, sex, weight, and electrophysiological and electroanatomical properties are extensively studied with compelling evidence. For instance, Laredo et al. (2018) found that multiple structural, electrophysiologic, ionic, and molecular changes observed during atrial fibrillation (AF) are more prevalent with older age, while younger age appears to be associated with protective properties against AF risks. Magnussen et al. (2017) investigated sex-specific differences in AF epidemiology and identified variations in prevalence, incidence, and risk factors, emphasizing the necessity for sex-specific prevention strategies. Siddiqi et al. (2022) further explored the contribution of body size factors to sex differences and concluded that sex differences in body size significantly influence the protective association between female sex and AF. Additionally, Johansson et al. (2020) discovered positive associations between height, weight, body mass index, and body surface with the risk of incident AF in both men and women. A recent study by Westaby et al. (2023) demonstrated that cardiac anatomical properties are associated with sex, age, and body measurements, with sex being the most discriminatory factor, specifically noting that the female heart exhibits smaller wall thickness, chambers, and valves.

The treatment of cardiovascular diseases is complex and requires a more targeted, patient-specific approach (Correa et al., 2020). In the realm of personalized medicine, increasing precision in treatment remains an ongoing endeavor, with complexities and costs escalating as efforts focus on pinpointing underlying molecular and cellular level differences. Consequently, identifying personalized cues in tissue-level recordings becomes paramount for individualized medical treatment.

This study aims to investigate the analysis of electrophysiological signals for predicting patient weight from intracardiac electrograms. A recent similar study by Alagoz (2024) explored the prediction of age from intracardiac electrograms. In this study, the state-of-the-art time series classification algorithm, Minirocket, was employed alongside the popular machine learning algorithm XGBoost. Minirocket consistently demonstrates superior performance compared to XGBoost across all class number scenarios and across all evaluation metrics, such as accuracy, F1 score, and AUC values, achieving scores of approximately 0.96. Conversely, XGBoost shows signs of overfitting, particularly noticeable in scenarios with higher class imbalance. Tuning probability thresholds for classes could potentially mitigate this issue. Additionally, XGBoost’s performance improves with reduced bin numbers, underscoring the importance of balanced classes. This study offers novel insights into the predictive capabilities of these algorithms and their implications for personalized medicine and remote health monitoring.

## MATERIALS AND METHODS

### Patient Population

The dataset utilized in this study was generously provided by Rodrigo et al. in (2022). This dataset consisted of a meticulously curated patient cohort sourced from the COMPARE registry (ClinicalTrials.gov Identifier: NCT02997254), comprising individuals diagnosed with AF who were prospectively enrolled during ablation procedures for symptomatic AF unresponsive to at least one anti-arrhythmic medication. Within this registry, intracardiac EGM recordings were performed on each patient using multipolar 64-pole basket catheters. A comprehensive review of the registry was conducted by a panel of three cardiac electrophysiologists, who meticulously classified each EGM tracing as either AF or Atrial Tachycardia (AT).

For this study, a systematic selection of patients from the registry was carried out to establish a balanced dataset of intracardiac recordings, comprising both AF (N = 22) and AT (N = 22) cases. It is important to note that all participants in this study provided written informed consent, in accordance with protocols approved by the Human Research Protection Program. A summary of patient demographics is provided in Table 1.

**Table 1.** The demographic information of the patients participating in the study.

|  | Patients (all) | Patients (AF) | Patients (AT) |
| --- | --- | --- | --- |
| Patient count | 44 | 22 | 22 |
| Age (in years) | 59.2±11.5 | 59.7±12 | 58.7±11.1 |
| Female | 14(31%) | 7(31%) | 7(31%) |
| Weight | 94.9±17.9 | 97.6±17.8 | 92.1±17.8 |

### Electrogram Collection

The dataset utilized in this study was a subset of the dataset originally provided by Rodrigo et al. (2022). Specifically, the analysis included N=2,817 EGM signals, with N=1,738 classified as AF and N=1,079 as AT. The procedure for EGM collection and pre-processing is outlined as follows.

During the electrophysiology study, a 64-pole basket catheter (Abbott, Menlo Park, CA) with electrodes sized at 2 mm and inter-electrode spacing set at 5 mm along the spline was used. Mapping was performed comprehensively across both the right and left atria. Unipolar EGMs spanning 60 seconds were extracted from the electrophysiological recorder (Prucka, GE Marquette, Milwaukee, WI; Bard Electrophysiology, Billerica, MA) and filtered within the frequency range of 0.05–500 Hz.

For analysis purposes, unipolar EGMs with a duration of 4 seconds, approximately equivalent to 20 cycles of AF or AT, were utilized. This duration aligns with the standard practice of examining EGM sequences in the frequency domain, with prolonged durations offering minimal enhancements in rhythm identification. The original EGMs, recorded at a sample frequency of 1 kHz (Bard) or 977 Hz (Prucka), were uniformly resampled to facilitate comparative analyses across datasets. To reduce dimensionality and considering that the physiological content of AF and AT EGMs lies below 200 Hz (Rodrigo et al., 2021), the EGMs were downsampled to 400 Hz utilizing a 200 Hz anti-aliasing filter. Ventricular artifacts were mitigated by subtracting the mean QRS complex, which was identified in three orthogonal ECG leads using a voltage threshold, and then averaged across a 1-minute period (Alhusseini et al., 2020).

### Specifying Classes

Since weight represents a continuous target variable, it undergoes transformation into an interval variable through discretization using a uniform strategy. This ensures that all bins possess identical widths. Subsequently, based on a predetermined number of bins, each weight value is assigned to the corresponding interval. For example, within an 8-bin setup with specified bin edges (refer to Table 2), a weight value of 82 falls within the interval [80,89). The associated class label is designated as “<89”, indicating that the value 82 is classified under the “<89” category. Throughout the study, various bin numbers corresponding to distinct numbers of classes are examined. Specifically, experiments are conducted using 8, 6, and 4 bins. The distributions of class membership counts are depicted in Figure 1. It is noteworthy that the balance among classes diminishes as the number of classes increases.

**Table 2.** Classification results.

| Model | Accuracy | F1 | AUC |
| --- | --- | --- | --- |
| *8-bin with edges:* [64, 72, 80, 89, 97, 106, 114, 122, 131] |  |  |  |
| Minirocket | 0.9605 ± 0.0069 | 0.9620 ± 0.0074 | 0.9777 ± 0.0045 |
| XGBoost | 0.7876 ± 0.0108 | 0.7592 ± 0.0163 | 0.9648 ± 0.0022 |
| *6-bin with edges:* [64, 75, 86, 97, 108, 120, 131] |  |  |  |
| Minirocket | 0.9629 ± 0.0094 | 0.9654 ± 0.0088 | 0.9785 ± 0.0055 |
| XGBoost | 0.8046 ± 0.0165 | 0.8053 ± 0.0182 | 0.9679 ± 0.0056 |
| *4-bin with edges:* [64, 80, 97, 114, 131] |  |  |  |
| Minirocket | 0.9493 ± 0.0052 | 0.9490 ± 0.0051 | 0.9654 ± 0.0035 |
| XGBoost | 0.8188 ± 0.0175 | 0.8175 ± 0.0174 | 0.9528 ± 0.0045 |

**Figure 1.**
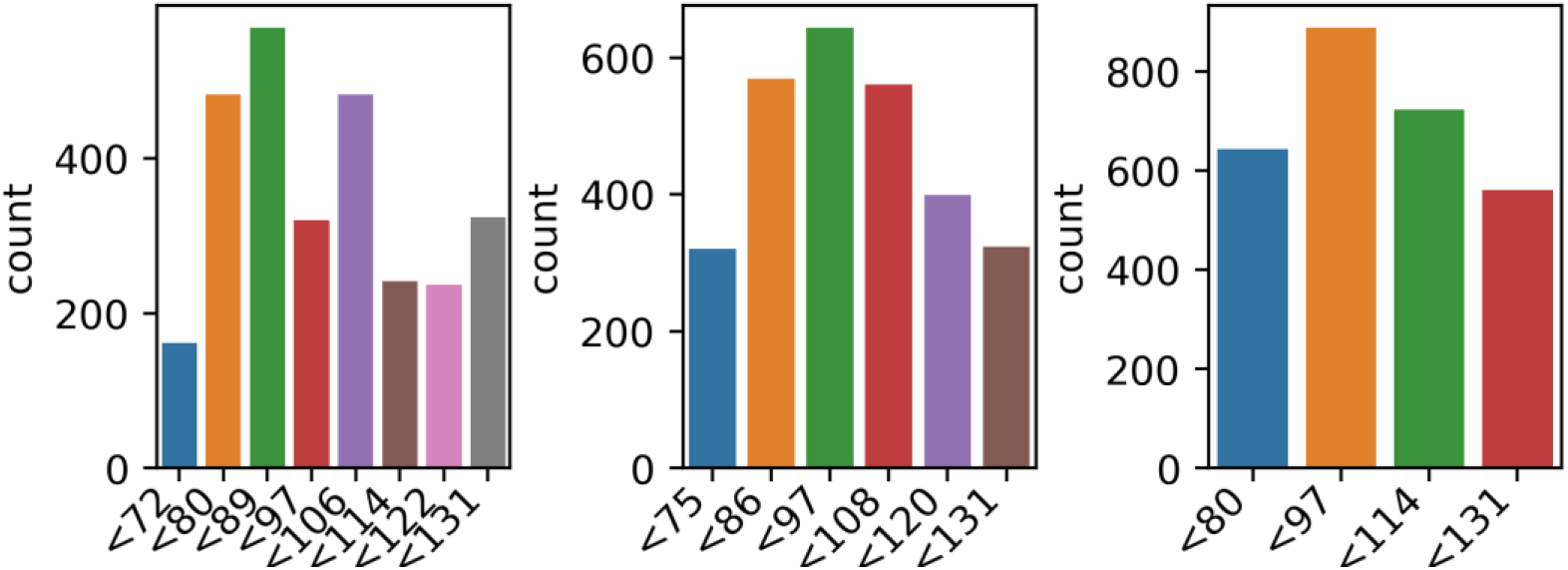
Counts of weight class members when bin size is chosen 8 (right), 6 (middle), 4(left).

### MiniRocket

MiniRocket is a feature extraction method proposed for tasks involving time series classification. Dempster et al. (2020) developed it with the aim of efficiently extracting features from time series data. MiniRocket applies random convolutional kernels to the input time series data and calculates statistical moments from the resulting random projections. These statistical moments serve as features that can be input into a classification algorithm. MiniRocket is recognized for its speed and scalability, making it suitable for large-scale time series classification tasks.

### Extreme Gradient Boosting

XGBoost is a widely used and powerful implementation of gradient boosting algorithms (Chen and Guestrin, 2016). It is popular in both machine learning competitions and real-world applications due to its efficiency, speed, and high performance. XGBoost constructs an ensemble of weak decision trees sequentially, with each tree correcting errors made by the preceding trees. It incorporates regularization techniques to prevent overfitting and supports parallel processing for accelerated training. XGBoost is effective for both classification and regression tasks and is known for its robustness and accuracy.

### Performance Evaluation

To ensure robust cross-validation, Monte Carlo cross-validation is employed, involving random partitioning of the dataset into training and testing sets across multiple iterations. Various combinations of training and testing sizes are explored to comprehensively assess classifier performance. To ensure the reproducibility of training and testing sets, the shuffling seed for each iteration is determined by the iteration number.

#### Accuracy

This metric assesses the overall correctness of the classifier by determining the ratio of correctly predicted instances to the total instances, offering an intuitive assessment of its performance.

#### F1 Score

The F1 score is a commonly used metric for evaluating classifier performance in binary classification tasks. It calculates the harmonic mean of precision and recall, providing a balanced measure of the classifier’s ability to correctly identify positive instances while minimizing false positives and false negatives. In multi-class scenarios, the study adopts the macro-averaging approach, where the F1 score is independently computed for each class and then averaged across all classes, assigning equal weight to each class.

#### AUC

The AUC is a metric used to evaluate the performance of binary classification models. It represents the area under the Receiver Operating Characteristic curve, which plots the true positive rate (sensitivity) against the false positive rate (1 - specificity) at various threshold settings. A higher AUC value indicates superior discrimination ability of the model. In multi-class classification tasks, the study adopts the One-vs-One (OvO) approach, where pairwise comparisons are made between all pairs of classes. For each pair of classes, a binary classifier is trained to distinguish between the instances of those two classes. The AUC is then calculated for each pair of classes, and the average AUC across all pairs is computed as the final multi-class AUC.

The experiments are conducted on a system equipped with an Intel(R) Core(TM) i7-5820K CPU @ 3.30GHz x 11 and 16 GB of RAM, utilizing the Python programming language for computations.

## RESULTS AND DISCUSSIONS

This study focuses on predicting the weight of a patient using intracardiac EGMs measured over a 4-second interval, employing Minirocket and XGBoost algorithms for prediction.

Table 2 presents the classification results. Minirocket consistently outperforms XGBoost with accuracy, F1 scores, and AUC values hovering around 0.96. On the other hand, XGBoost demonstrates accuracy and F1 scores around 0.80, while achieving AUC values of approximately 0.96. This discrepancy suggests that XGBoost may be overfitting, potentially remediable through tuning probability thresholds for classes. Additionally, XGBoost’s performance improves with a reduction in bin numbers, leading to more balanced classes. The decrease in class imbalance with fewer bins indicates a possible cause for XGBoost’s overfitting.

Contrastingly, Minirocket displays robustness to class imbalance, maintaining consistent performance around 0.96 across varying class numbers. However, in the case of 4-bin classification, F1 and accuracy values slightly drop to around 0.95. It is noteworthy that Minirocket performs better with a greater number of classes and increased class imbalance, which is unexpected.

Further insights can be gleaned from the confusion matrices depicted in Figure 2, particularly for the 8-class scenario. XGBoost exhibits lower precision and recall for classes with relatively fewer members, as indicated in Figure 1. This highlights XGBoost’s susceptibility to overfitting due to class imbalance. Conversely, Minirocket demonstrates robust performance, achieving precision and recall values of 1.0 for class “<72”, while XGBoost’s values are 0.92 for the same class.

**Figure 2.**
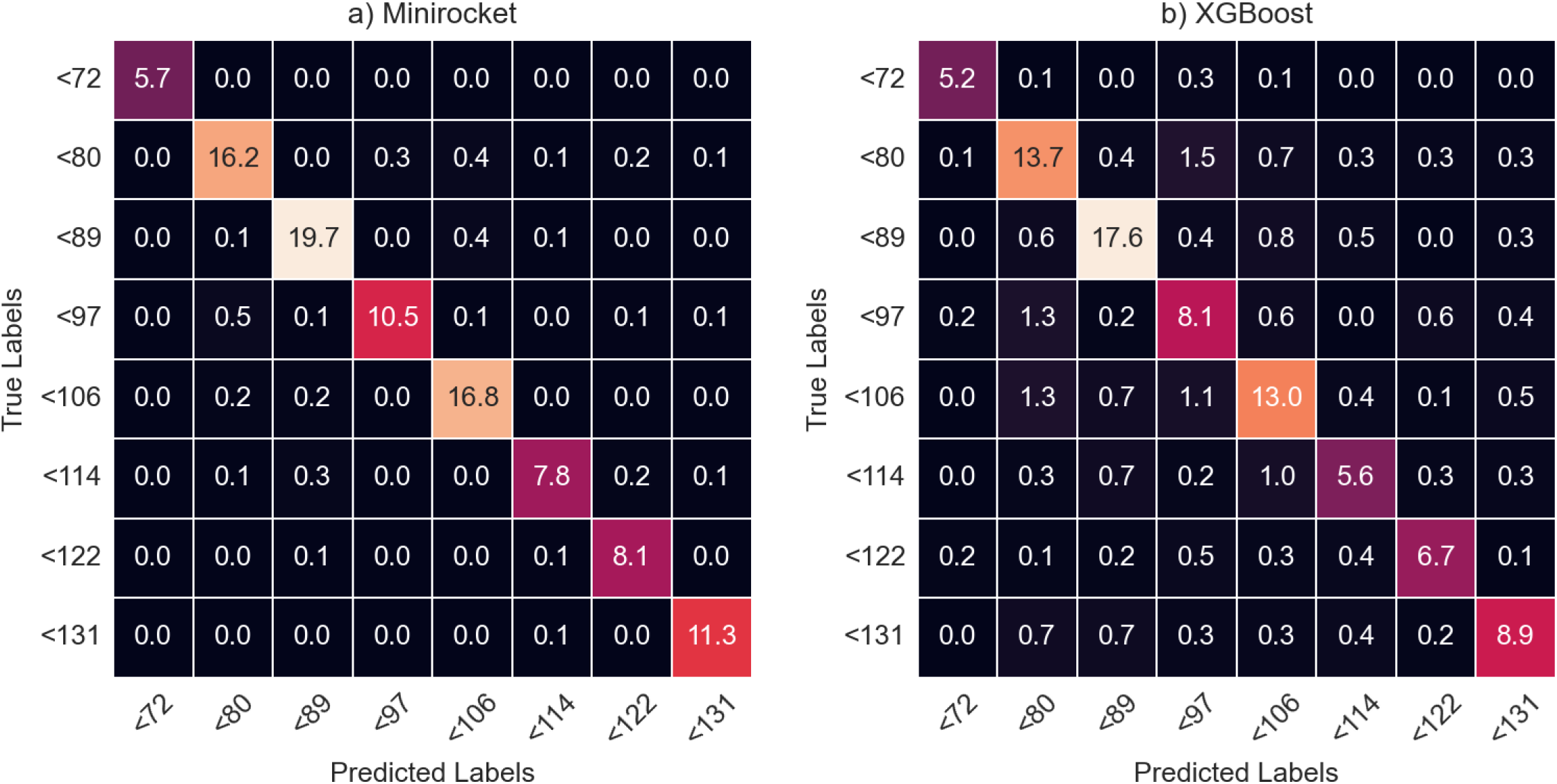
Confusion matrices resulting from Monte Carlo cross-validation runs. Cell values denote the percentages of total number of test instances used in all runs.

In a broader context, this study introduces a novel approach enabling the estimation of patient demographics from electrophysiological signals, a pioneering effort in the literature. The significance of this prediction lies in several aspects. Firstly, in conventional medical settings, patient information is typically gathered through personal history assessment before obtaining electrophysiological measurements, which then inform diagnostic or therapeutic actions. Hence, accurate estimation of patient demographics is crucial. Secondly, in scenarios such as urgent or ambulatory cases where patient information may not be readily accessible, precise estimation of patient demographics becomes indispensable. Thirdly, amidst the rapid development of wearable technology and remote data collection, the assessment of auxiliary data aids in enhancing remote health monitoring. Lastly, deriving patient-specific cues from signals obtained directly from the patient holds substantial implications for personalized medicine.

## CONCLUSIONS

In conclusion, this study demonstrates the efficacy of Minirocket and XGBoost algorithms in predicting patient weight using intracardiac EGMs. Minirocket consistently outperforms XGBoost across various metrics, showcasing its robustness to class imbalance and superior performance in scenarios with a greater number of classes.

The findings suggest that XGBoost may be overfitting, particularly evident in scenarios with higher class imbalance. However, this overfitting can potentially be mitigated through tuning probability thresholds for classes. Furthermore, XGBoost’s performance improves with a reduction in bin numbers, indicating the importance of balanced classes in enhancing algorithm performance.

On the other hand, Minirocket maintains consistent performance across different class numbers, highlighting its resilience to class imbalance. Nevertheless, a slight drop in performance is observed in the 4-bin classification scenario.

The analysis of confusion matrices provides deeper insights into the performance of both algorithms, with XGBoost displaying lower precision and recall for classes with fewer members compared to Minirocket.

Overall, this study introduces a novel approach for estimating patient demographics from electrophysiological signals, presenting significant implications for personalized medicine, remote health monitoring, and scenarios where patient information may not be readily accessible. Further research could focus on refining algorithms to address overfitting issues and exploring additional features for improved prediction accuracy.

## Data Availability

All data produced in the present study are available upon reasonable request to the authors.

